# Development and Evaluation of an AI System for COVID-19 Diagnosis

**DOI:** 10.1101/2020.03.20.20039834

**Authors:** Cheng Jin, Weixiang Chen, Yukun Cao, Zhanwei Xu, Zimeng Tan, Xin Zhang, Lei Deng, Chuansheng Zheng, Jie Zhou, Heshui Shi, Jianjiang Feng

## Abstract

Early detection of COVID-19 based on chest CT will enable timely treatment of patients and help control the spread of the disease. With rapid spreading of COVID-19 in many countries, however, CT volumes of suspicious patients are increasing at a speed much faster than the availability of human experts. We proposed an artificial intelligence (AI) system for fast COVID-19 detection and performed extensive statistical analysis of CTs of COVID-19 based on the AI system. We developed and evaluated our system on a large dataset with more than 10 thousand CT volumes from COVID-19, influenza-A/B, non-viral community acquired pneumonia (CAP) and non-pneumonia subjects. In such a difficult multi-class diagnosis task, our deep convolutional neural network-based system is able to achieve an area under the receiver operating characteristic curve (AUC) of 97.17%, a sensitivity of 90.19%, and a specificity of 95.76% for COVID-19 on internal test cohort of 3,203 scans and AUC of 97.77% on the publicly available CC-CCII database with 1,943 test samples. In a reader study involving five radiologists, the AI system outperforms all of radiologists in more challenging tasks at a speed of two orders of magnitude above them. Diagnosis performance of chest x-ray (CXR) is compared. Detailed interpretation of deep network is also performed to relate AI results with CT findings. The code is available at https://github.com/ChenWWWeixiang/diagnosis_covid19.

---

The COVID-19 pandemic^1^ has infected more than 6 million people worldwide, killed more than 360 thousand people and is still spreading rapidly world-wide. It is important to detect COVID-19 as quickly and accurately as possible for controlling the spread of the disease and treating patients. Even though reverse transcription-polymerase chain reaction (RT-PCR) is still ground truth of COVID-19 diagnosis, the sensitivity of RT-PCR is not high enough for low viral load present in test specimens or laboratory error^2^, and kits of RT-PCR are in short of supply in many areas^3^.

As a result, some contourites used chest imaging such as chest CT or chest x-ray (CXR) as first-line investigation and patient management tools^4,5^. Chest imaging, especially CT, can show early lesions in the lung and, if diagnosed by experienced radiologists, can achieve high sensitivity^3^. In addition, CT scans are widely available and economic. At present, the diagnosis of chest CT depends on the radiologists, which has some problems. Firstly, chest CT contains hundreds of slices, which takes a long time to diagnose. Secondly, COVID-19, as a new lung disease, has similar manifestations with various types of pneumonia^6^. Radiologists need to accumulate a lot of CT diagnostic experience to achieve a high diagnostic performance. In some outbreak areas, many suspected patients are in urgent need of diagnosis and proper treatment, and many CT scans are performed every day. There is an urgent shortage of radiologists with high diagnosis performance for COVID-19.

Artificial intelligence (AI) may be the unique preparation to take up this challenge. Powered by large labeled datasets^7^ and modern GPUs, AI, especially deep learning technique^8^, has achieved excellent performance in several computer vision tasks such as image classification^9^ and object detection^10^. Recent research shows that AI algorithms can even achieve or exceed the performance of human experts in certain medical image diagnosis tasks^11-15^. The AI diagnosis algorithm also has the advantages of high efficiency, high repeatability and easy large-scale deployment. The current outbreak of COVID-19 is worldwide, and the shortage of specialist radiologists threatens the availability and adequacy of screening services for COVID-19 in affected areas. By deploying AI diagnosis algorithms, suspicious patients everywhere, especially in developing countries, will have equal access to right diagnosis, timely treatment and isolation.

There are already some published studies on CT based COVID-19 diagnosis systems^16,17^. Here we briefly review several representative studies employing relatively large datasets. Zhang *et al*.^18^ developed COVID-19 diagnosis system on a database consisting 4,154 patients and it can differentiate COVID-19 from other common pneumonia and normal healthy with AUC of 0.9797. In their system, the classification was based on lesion segmentation result and the lesion segmentation DICE-index was about 66.2%, which is not an accurate representation of lesions. Another drawback is manual annotation of training segmentation masks is a very expensive procedure. Li *et al*.^19^ developed an AI system and yielded AUC of 0.96 for COVID-19 detection on dataset consisting of 3,322 subjects including COVID-19, CAP and healthy people. Their system extracted features on slices and fuse them into volume-level which increases much memory demand while without extracting more informative 3D features. Several slice level diagnosis methods^20-22^ were proposed which were quite similar to Li *et al*.’s work. Some AI systems employed 3D convolution neural networks, but they considered only the relatively simple two-category classification ^23,24^ There are also a few COVID-19 detection systems using CXR^25^, but the number of subjects with COVID-19 in these studies is much smaller than the studies using CT and no study has quantitively compared performances of CXR and CT using paired data.

Our study has some important differences from previous studies. We constructed clinically representative large-scale datasets with 10,250 CT scans from three centers in China and three publicly available databases, which is much larger than previous studies. To understand relative performances of CT and CXR for detecting COVID-19, we developed both CT-based and CXR-based diagnosis systems and tested them using paired data, which has not been studied before. We compared the diagnostic performance of our CT-based diagnosis system with that of five radiologists in reader studies, and the results showed the performance of this system is higher than that of experienced radiologists. In addition, based on prediction score on every slice of CT volume, we can locate the lesion areas in COVID-19 patients and performed a statistical study of different subsets of patients. The specific phenotypic basis of the diagnosis output was also traced by an interpretation network and radiomics analysis was applied to understand the imaging characteristics of COVID-19.

## Result

### Datasets for System Development and Evaluation

We developed and evaluated a deep learning-based COVID-19 diagnosis system, using multiclass multi-center data, which includes 10,250 CT scans from 7,917 subjects consisting of COVID-19, CAP, influenza and non-pneumonia. CAP subjects included in our database were all non-viral CAP. Data were collected in three different centers in Wuhan, and from three publicly available databases, LIDC-IDRI^26^, Tianchi-Alibaba^27^, and CC-CCII^18^ (detailly described in **Extended Data Table 1** and **Methods**).

Due to multi-stage CT scans of the same person might be similar, the cohort division was performed on subjects with no overlapping subjects in different sub-cohorts. Except CC-CCII, all remaining data was divided into two independent parts, a training cohort of 2,688 subjects and a test cohort of 2,609 subjects. Three reader study cohorts were randomly chosen from test cohort with respectively 100, 100 and 50 subjects for three tasks, classifying pneumonia from healthy, classifying COVID-19 from CAP, and classifying COVID-19 from influenza. Only image slices of JPEG format rather than raw volumes are available on CC-CCII databases, therefore we had to train and evaluate specially for CC-CCII. CC-CCII cohort contains 2,539 subjects which was divided into a training cohort of 1,269 subjects and a test cohort of 1,270 subjects (detailly described in **Methods**).

### Construction of the AI System for COVID-19 Diagnosis

We proposed a deep-learning based AI system for COVID-19 diagnosis, which directly takes CT data as input, performs lung segmentation, COVID-19 diagnosis and COVID-infectious slices locating. In addition, we hope that the diagnosis results of AI system can be quantitatively explained in the original image to alleviate the drawback of deep neural networks as a black box. The system consists of five key parts (**Figure 1 A**), (1) lung segmentation network, (2) slice diagnosis network, (3) COVID-infectious slice locating network, (4) visualization module for interpreting the attentional region of deep networks, and (5) image phenotype analysis module for explaining the features of the attentional region.

**Figure 1.**
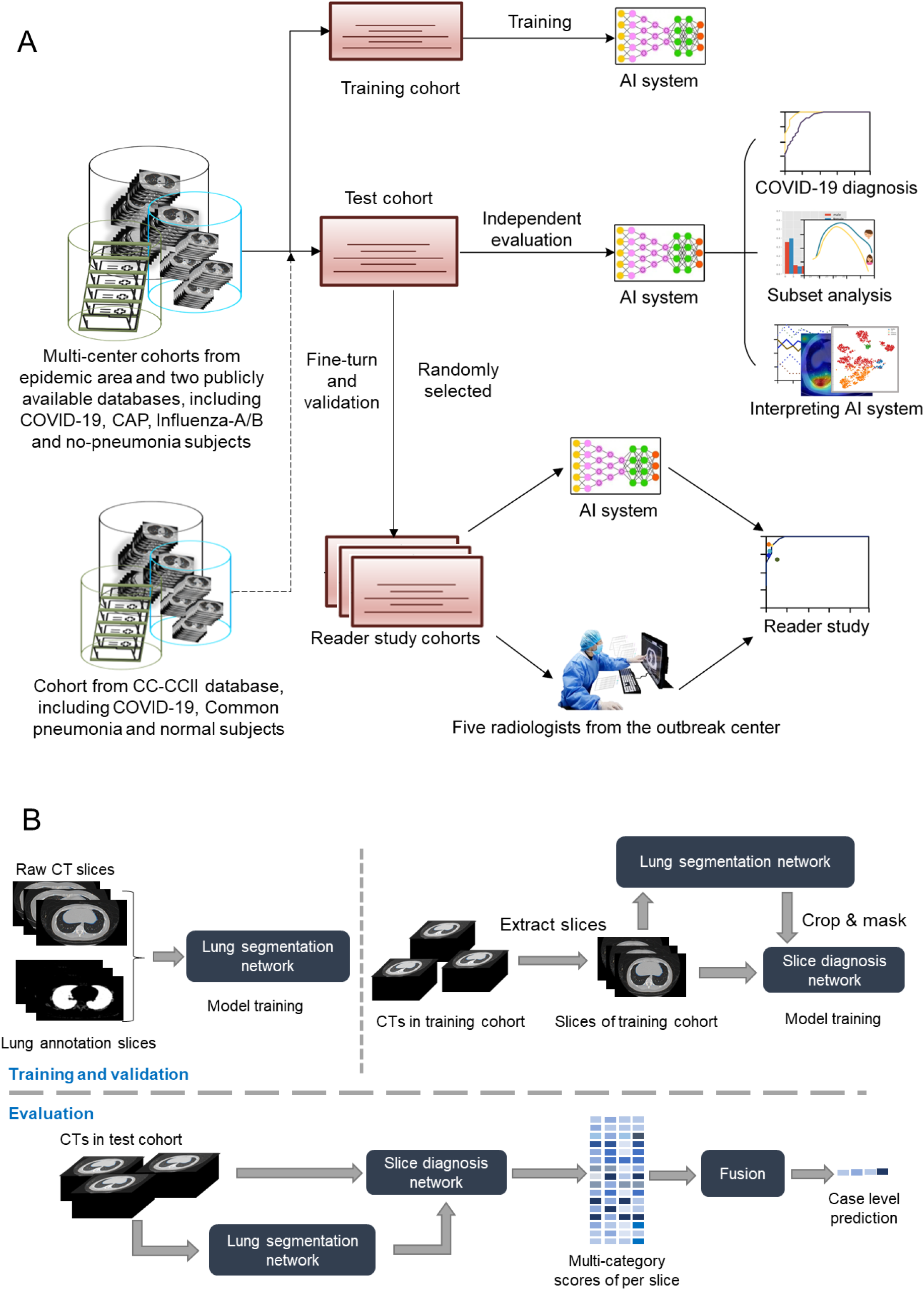
Workflows of the whole study and the proposed AI system. A. Workflow of the whole study. B. Construction and usage of the AI system.

The workflow of deep-learning based diagnosis model is shown in **Figure 1 B**. CT volumes were divided into different cohorts. Then after slice level training, our model can accurately classify input slices into four categories, including non-pneumonia, CAP, influenza-A or B and COVID-19. A task-specific fusion module was proposed to our model fused slice results into case level diagnosis according to different diagnosis tasks, as a result the network can be used in different tasks without retraining. The model was implemented in 2D not only because it is easier to train within memory limit of common GPUs (usually 11G), but also because slice-level diagnosis can be used for COVID-infectious slice locating. Other modules of our system are described in **supplementary methods**.

### Performances of Diagnosis

The trained AI system was evaluated on the test cohort. We used the receiver operating characteristic (ROC) curves to evaluate the diagnostic accuracy. On the test cohort, the ROC curve (**Figure 2 A**) showed AUC of four categories were respectively 0.9752 (for non-pneumonia), 0.9804 (for CAP), 0.9885 (for influenza) and 0.9745 (for COVID-19). Besides, sensitivity and specificity of COVID-19 were 0.9019 and 0.9576 (**Extended Data Table 2 A**). Our system showed great generalization ability with 0.9778 AUC for COVID-19 in publicly available database CC-CCII (**Figure 2 B, Extended Data Table 2 B**). The confusion matrix of four categories and PR curves of diagnosis of COVID-19 are shown on **Extended Data Figure 1 C**. The decision curve analysis (DCA)s for the AI system are presented in **Extended Data Figure 1 D-F**, which indicated that the AI system added benefit when the threshold was within wide ranges of 0.03-0.87, 0.04-0.90, 0.20-1.00 separately for COVID-19 diagnosis with negatives from CAP, non-pneumonia and influenza. Confusion matrixes of test cohort and CC-CCII test cohort are show in **Extended Data Figure 1 A, B**.

**Figure 2.**
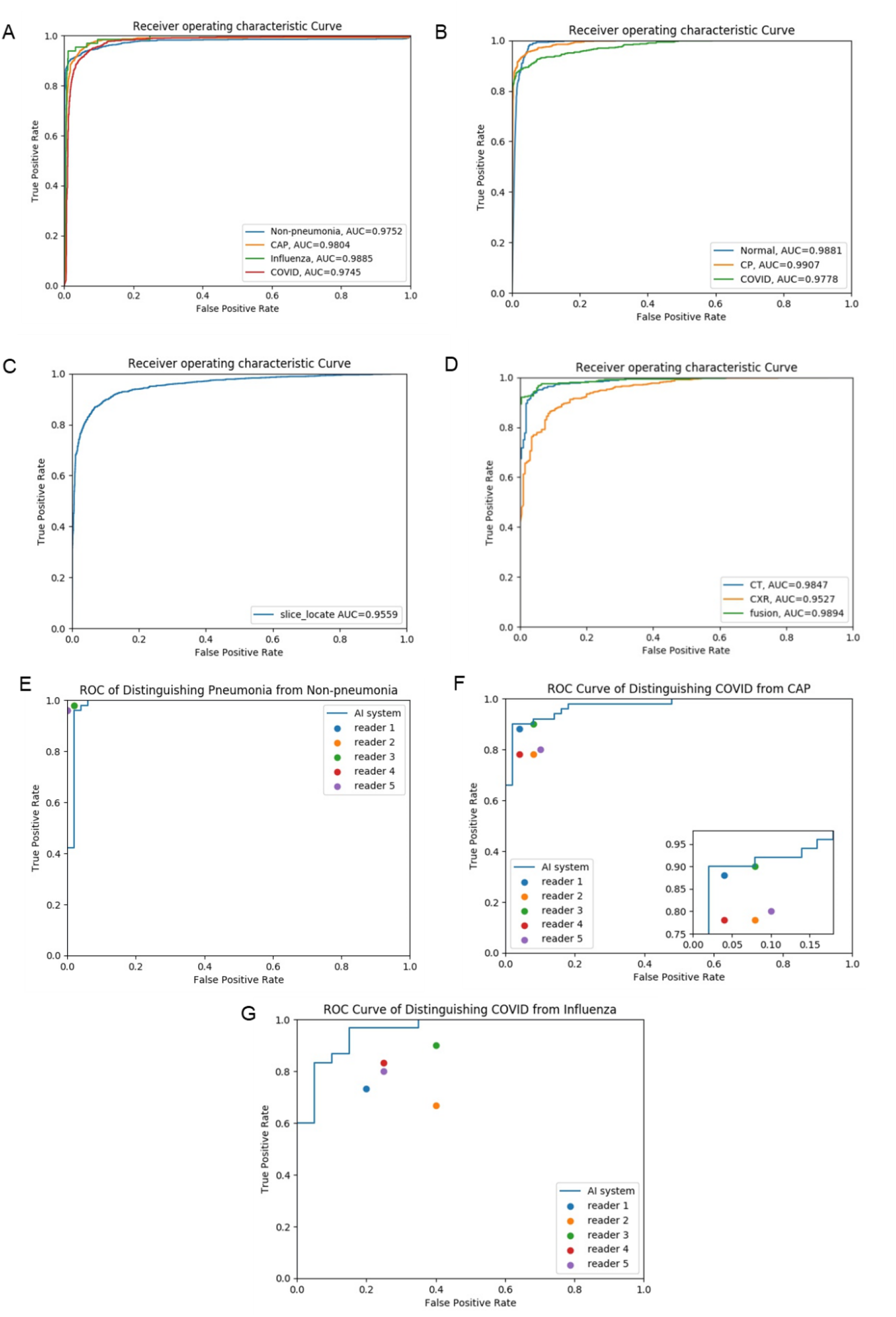
Receiver Operating Curves of the AI system. **A**. ROC curves of AI system on our test cohort. **B**. ROC curves of AI system on CC-CCII test cohort. **C**. ROC curve of AI system on COVID-infectious locating. **D**. ROC curves of CT-based AI system and CXR-based AI system on sub-cohort of test cohort which has paired CT and CXR data. **E**. ROC curve together with reader performances on pneumonia-or-non-pneumonia cohort. **F**. ROC curve together with reader performances on CAP-or-COVID cohort. **G**. ROC curve together with reader performances on Influenza- or-COVID cohort.

### Performances of COVID-Infectious Locating

COVID-infectious slice locating results are shown in **Figure 2 C**. Although with a same network structure as the slice diagnosis network, our experiments showed training COVID-infectious slice locating network using normal and abnormal slices from COVID-19 subjects led to a much better performance, with AUC of 0.9559, specificity of 0.9636, and sensitivity of 0.8009.

### Comparison of AI System to Radiologists

We conducted a reader study with five board-certified radiologists (Average of 8 years clinical experience, range 5-11 years, **Extended Data Table 3 A**). All readers were asked to read independently without other information about patients.

Different from Zhang *et al*.^18^, three different cohorts with different tasks were setup for reader study, which were pneumonia-or-non-pneumonia cohort, CAP-or-COVID-19 cohort and influenza-or-COVID-19 cohort (detailly described in **Methods)**. The separated tasks helped us to detailly analyze the COVID-19 distinguishing ability with different negative classes. ROC curves of the AI system in these tasks were presented to compare with the performances of human radiologists **(Figure 2 E, F, G, Extended Data Table 2 D)**. Only in the pneumonia-or-non-pneumonia cohort, the AI system performs slightly worse than radiologists. In CAP-or-COVID-19 and Influenza-or-COVID-19 cohorts, the AI system outperformed all human radiologists. **Extended Data Table 3 B, C** shows comparison of diagnostic error between the AI system and human readers.

**Extended Data Figure 2** shows some slices from error predictions of the AI system and human readers. Human readers tend to use some macro-level radiology features in diagnosis, so that the error predictions of human had nontypical presentation, while the AI system can overcome this. For example, the CAP in **Extended Data Figure 2 B** looks like typical ground-glass opacity (GGO) and COVID-19 in that figure has nontypical density decrease (blue arrow).

### Subset Analysis

For an in-depth understanding of the AI system and characteristics of different populations with COVID-19, we evaluated the AI system on subsets of test cohort divided by gender, age and stage. **Figure 3 A** shows the AUC of four categories in subsets separated by gender, age and stage. To understand the cause for different diagnosis performances, we analyzed the COVID-infectious slice locating results in different subsets (**Figure 3 B**). Subjects from LIDC-IDRI and Tianchi-Alibaba had been anonymized so that the analysis were not done on them.

**Figure 3.**
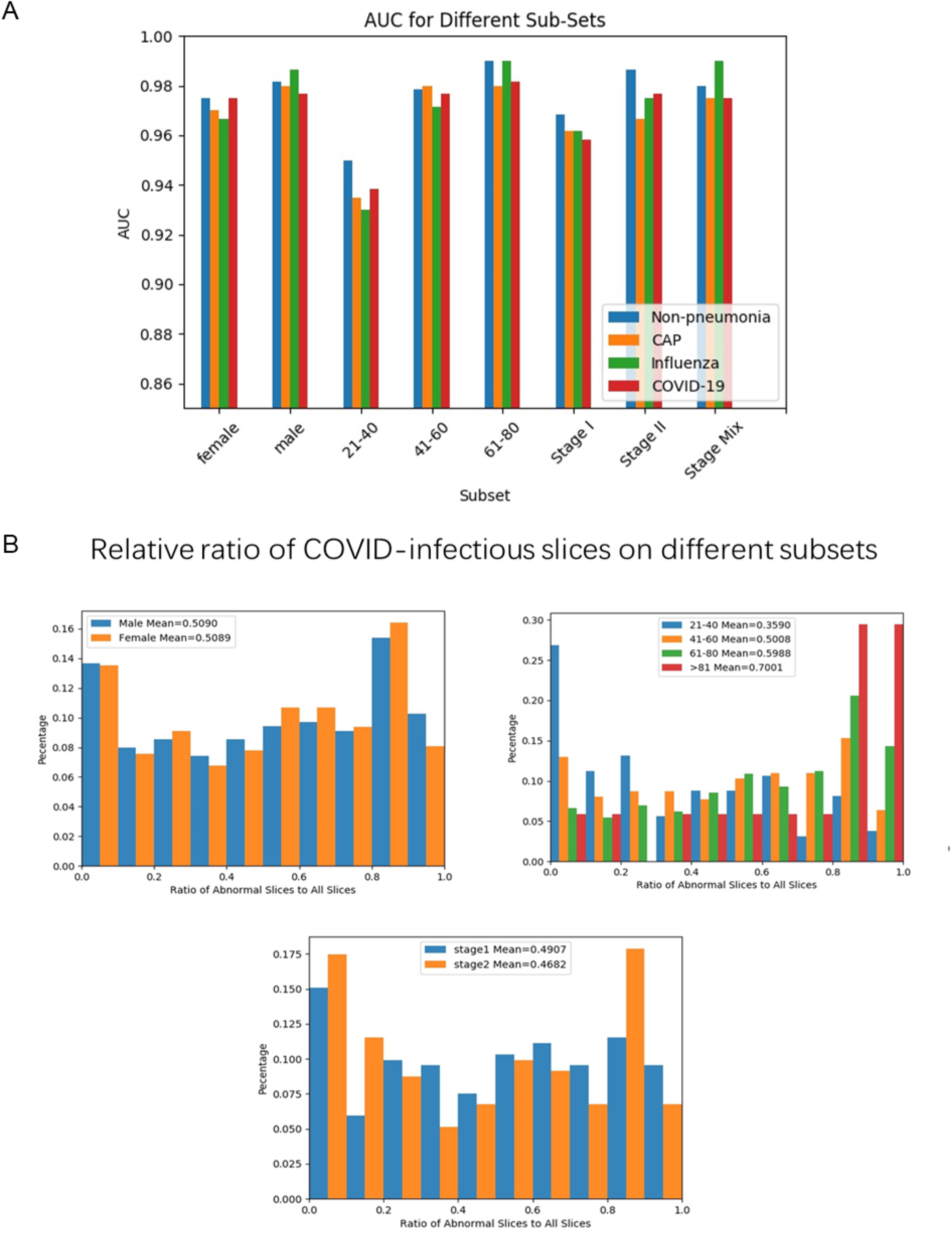
Statistics on different subsets of subjects in test cohort. **A**. AUC scores for each category on different subsets. **B**. Distribution of ratio of COVID-infectious slices on different subsets of COVID-19 patients.

A subset of the patients in the database have multi-stage CTs. We compared the diagnostic performance of stage I and stage II and fusion of them. The experiment suggested that the performance of the AI system is independent of the progress of the disease because of no significant qualitative differences between performances of different stages. We did not test more complex fusion methods which may overestimate the performance since most non-pneumonia and CAP subjects have only one stage CT.

A subset of COVID-19 and CAP subjects in the database have localizer scans along with CT scans. The localizer scans of CTs are very similar to CXR but typically noisier. We used this subset to study performances of CT versus CXR. We developed a CNN based classification algorithm to discriminate COVID-19 from CAP using these localizer scans (detailly described in **Methods** and supplementary method). Experiments on subjects with both types of data showed that CT-based system performed significantly better (**Figure 2 D, Extended Data Table 2 C**). Representative examples are given in **Extended Data Figure 3**.

### Interpreting the AI System

After proper training of the deep network, Guided gradient-weighted Class Activation Mapping (Guided Grad-CAM) ^28^ was exploited to explain the “black box” system and extract attentional areas which is connected to the back end of the diagnostic model (Supplementary Figure 4). **Figure 4 A** shows some representative subjects for the visualization of Guided Grad-CAM to determine the attentional regions for each category. We used t-SNE^29^ to map 2048-dimensional deep features to 2-dimensional plane (**Figure 4 B**) and the result showed that our model extracted powerful features to separate different categories in latent space.

**Figure. 4.**
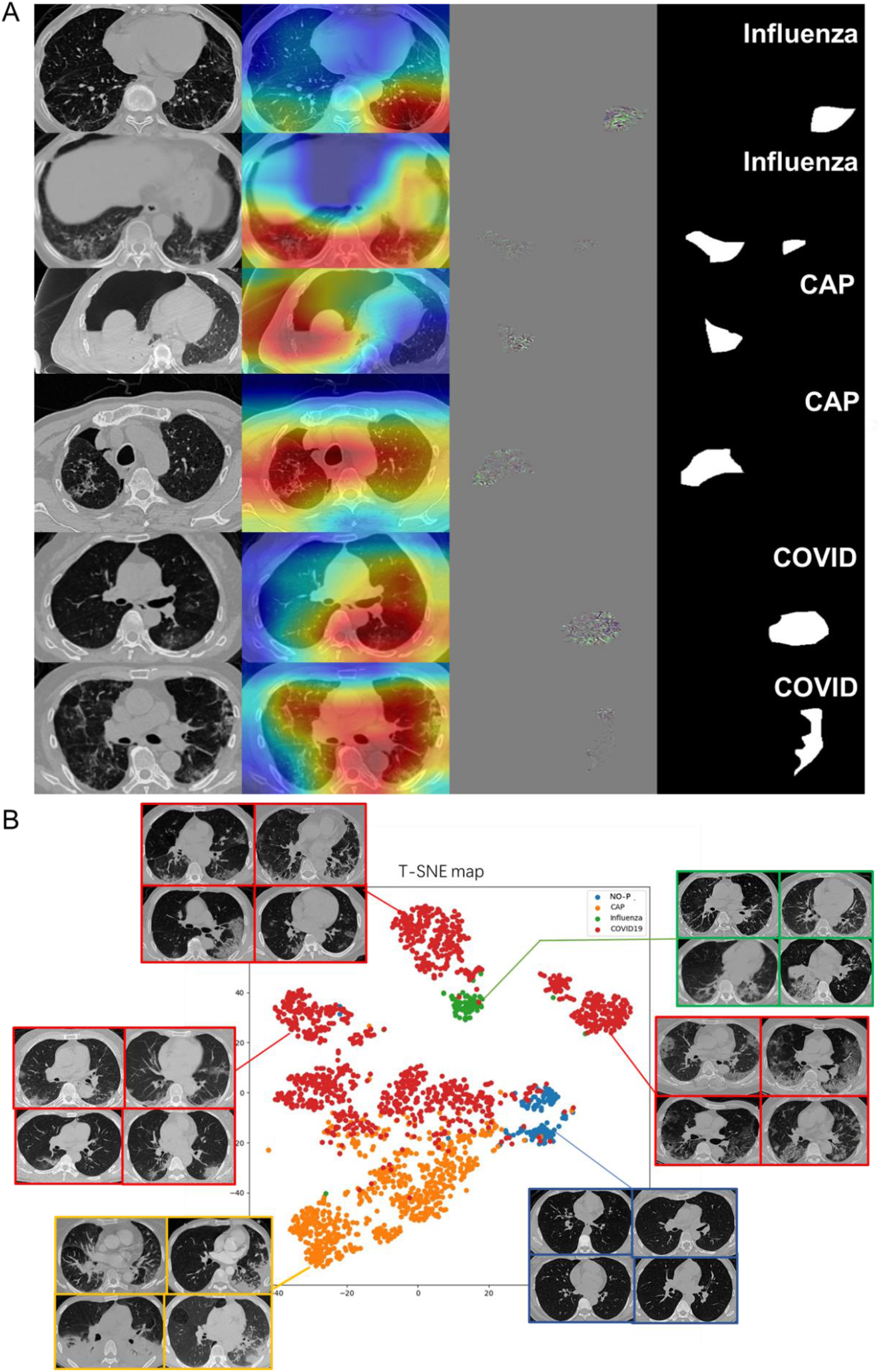
Interpreting the AI System. **A**. Visualizing feature maps via Grad-CAM. **Notes:** Three representative subjects for the visualization of AI diagnosis. From left to right: Original CT image; coarse-resolution attentional regions overlaid on CT image; high-resolution attentional regions with fine granularity; binarized maps of region of attention obtained from Guided Grad-CAM. **B**. Visualizing features via t-SNE

We performed radiomics^30^ feature extraction on these attentional regions, and obtained a total of 665-dimensional imaging features. The Least Absolute Shrinkage and Selection Algorithm (LASSO) were used to find the most discriminative 12 features in distinguishing COVID-19 from other pneumonia. Three additional features were also extracted for the attentional regions, distance feature, 2D margin fractal dimension, and 3D grayscale mesh fractal dimension (Phenotype Feature Extraction in **Methods**). The statistics of these features were consistent to previous literature^31^ on the pathogenesis and morphology of COVID-19. The selected features were used to explain the imaging characteristics in CT (Phenotype Feature Analysis in **Methods**) and t-test and Kolmogorov-Smirnov (KS) test were performed to statistically analysis those features (**Extended Data Figure 4-6**).

## Discussion

In this study, we developed an AI system for diagnosis of COVID-19, CAP, influenza and non-pneumonia. The system showed good sensitivity (0.8703), specificity (0.9660) and AUC (0.9745) for COVID-19 in test cohort. Furthermore, in the reader study, the diagnostic accuracy of the AI system outperformed experienced radiologists in two tasks from the outbreak center, with AUC of 0.9869, 0.9727, 0.9585 separately for pneumonia-or-non-pneumonia, CAP-or-COVID-19 and influenza-or-COVID-19 tasks. In the reader study, the average reading time of radiologists was 6.5 min, while that of AI system was 2.73 s, which can significantly improve the productivity of radiologists. Only in pneumonia-or-non-pneumonia cohort, the AI system worked slightly worse than human readers. In those more challenging tasks, the AI system worked better than human readers. For diagnosis between CAP and COVID-19, when the AI system misclassify, the radiologists were also wrong in 37.5% (3/8) of subjects (**Extended Data Table 3 B**), indicating that the diagnosis of these cases is challenging. And for influenza-A/B, that number is 50% (3/6). Meanwhile, we found that 88.5% (23/26) and 86.9% (20/23) of errors made by radiologists in those two tasks were correctly classified by the AI system. It means that the AI system can be used as an effective independent reader to provide reference suggestions. Besides, the AI system can be adapted to different requirements. According to a highly sensitive setting, it can screen out suspicious patients for confirmation by radiologists; with a high specific setting, it can warn possible diagnosis errors made by radiologists.

To further understand the performance of the AI system, we evaluated it on subsets divided by gender, age and stage (**Figure 3 A, B**), which can assist decision-making in different populations. According to **Figure 3 A, B**, the number of infectious slices was related to age, and the diagnostic performance was also related to age. We concluded that young people might have less infectious area resulting in lower diagnostic performance. The results on the subsets divided by gender showed little difference on infectious slices, but the average AUC for man was higher than that for women. This is consistent with the conclusion of Xiong. Q et al^32^. that women have higher antiviral immunity than men, so that it was harder for AI to find out diagnostic clues in CTs of women. The results of different stages showed that the performance of the AI system had little correlation with the stages of CT scans. Fusion of stages can slightly improve the performances.

CXR is also considered as a possible way to diagnose COVID-19. According to **Figure 2 D** and **Extended Data Figure 3**, CXR had diagnostic value though it was generally not better than CT. In the situation that patients might not hold breath well which brought in artifacts in CT, CXR worked better than CT (**Extended Data Figure 3 C**). By fusion with CXR using simple results level averaging, a slight benefit would be acquired compared to using CT only. **Extended Data Figure 3 E** showed an example that the result was corrected after fusion, while when CXR was in low quality, fusion can also bring in errors (e.g. **Extended Data Figure 3 F**). A better fusion method might help achieve better performance such as fusion in feature-level when training deep networks. As far as our survey, we are the first to compare CT and CXR performances in a paired cohort, which was fair to CT and CXR so that the comparation result was convincing. Importantly, the performance of CXR might be underestimated because localizer scans are in poor quality compared to normal CXR. Nevertheless, localizer might be the best possible data to compare CT against CXR, since there is currently no dataset with paired CT and normal CXR captured at very close time interval.

According to Grad-CAM, we found that the AI system focused on different regions on the types of pneumonias. For CAPs, it generally ignored GGO which might also occur in COVID-19 and focused on effusion and consolidation adjacent to the pleura. On the other hand, the AI system focused GGO rather than consolidation for most COVID-19 subjects. For influenza and COVID-19, Grad-CAM displays similar concerns, such as stripe consolidation and GGO, but the AI system is still able to distinguish the two correctly. We speculated that the AI system will focus on specific regions where other types of pneumonia may be rare. T-SNE clearly showed that deep feature provided by our AI system can divide different types of pneumonia into different clusters, as shown in **Figure 4 B**, especially COVID-19 subjects were mapped to more than one clusters. By visualizing the raw image of the feature points, COVID-19 was found to have several types of presentations (lefts, uppers and rights), which were enclosed by the red borders. Samples in left cluster of COVID-19 were most in early and mild stage which have small GGO with nearly round shape. Samples in right cluster had larger lesion and some of them had crazy paving patterns. Fibration and consolidation could be found in the upper cluster whose sizes of lesion were generally between lefts and rights. Although visualization by t-SNE was a conjecture for extracting features from the network, we can clearly find that patients of COVID-19 may be divided into different subclasses.

Further, we provided a visual interpretation of the system’s decision by performing a radiomics analysis to obtain diagnostically relevant phenotypic characteristics of the attentional regions that are fully traceable on the original CT image. By visualizing the diagnostic results and the phenotype analysis, we found that the spatial distribution of the attentional region, morphology and the texture within it are consistent with the characteristics of COVID-19 as reported in previous manual diagnosis studies^6,33^, and we can make pathophysiological and anatomical speculations on the viral infection process (see **Phenotype Feature Analysis** in **Methods**).

There are still some drawbacks and future works of this research. Firstly, collecting more data on more subtypes of pneumonias or other lung deceases is useful for explore AI system with higher diagnosis capability. Secondly, Guided Grad-CAM can only extract attentional region rather than lesion segmentation. Phenotype feature analysis would be better to be done on accurate segmentations. Finally, constructing a large dataset with linked CT and clinical information will enable more analysis.

Overall, the proposed AI system has been comprehensively validated on large multi-class dataset with diagnosis performance higher than human experts in diagnosing COVID-19. Unlike classical black-box deep learning approaches, by visualizing the AI system and applying radiomics analysis, it can decode effective representation of COVID-19 on CT imaging, and potentially lead to the discovery of new biomarkers. Radiologists could perform an individualized diagnosis of COVID-19 with the AI system, adding new driving force for fighting the global spread of outbreak.

## Data Availability

Three databases in our experiments are publicly available. LIDC-IDRI database can be accessed at https://wiki.cancerimagingarchive.net/display/Public/LIDC-IDRI. Tianchi-Alibaba database can be accessed at https://tianchi.aliyun.com/competition/entrance/231601/information. CC-CCII database can be accessed at http://ncov-ai.big.ac.cn/download.
The datasets from Wuhan Union Hospital, Western Campus of Wuhan Union Hospital, and Jianghan Mobile Cabin Hospital were used under the license of the current study and are not publicly available.

https://wiki.cancerimagingarchive.net/display/Public/LIDC-IDRI.

https://tianchi.aliyun.com/competition/entrance/231601/information

http://ncov-ai.big.ac.cn/download

## Methods

### Development and Validation Datasets

We collected data from both hospitals in Wuhan and publicly available databases.

Under institution review board (IRB) approval, we collected data from three centers in Wuhan, which are Wuhan Union Hospital, Western Campus of Wuhan Union Hospital, and Jianghan Mobile Cabin Hospital. In total, 4,260 CT scans (2,529 COVID-19 scans, 1,338 CAP scans, 135 influenza-A/B scans and 258 normal scans) from 3,177 subjects (1502 COVID-19 patients, 83 influenza-A/B patients, 1334 CAP patients except influenza, and 258 healthy subjects) were collected from multi-centers **(Extended Data Table 1)**. CT volumes of COVID-19 patients were collected from February 5th, 2020 to March 29th, 2020, and all these patients were confirmed as COVID-19 by RT-PCR. For subjects with three or more scans, we excluded the last scan since the last ones might be rehabilitative. Extended Data Table 1 B shows characteristics of multi-scan data after exclusion. CTs of heathy subjects are from physical examinations of Union Hospital from January 2nd, 2020 to February 2nd, 2020. All these subjects were PCR negative and no pneumonia signs were found in their CTs according to CT diagnosis report. CAP volumes were collected from January, 2019 to November, 2019. The CAP cases in our cohort were all non-viral pneumonias. Influenza-A/B volumes were collected from November, 2016 to November, 2019. All CAP and influenza subjects were retrospective and confirmed subjects which must not be COVID-19 according to study dates.

LIDC-IDRI and Tianchi-Alibaba are both databases for lung nodule detection with separately 1,009 and 1,200 scans available. All subjects of them suffered from benign or malignant lung nodules. Because nodules have totally different presentations, we setup a category “non-pneumonia” to cover both healthy subjects from Wuhan and subjects from LIDC-IDRI and Tianchi-Alibaba.

All above data was randomly divided into two independent parts with no overlapping subjects. The ratio of division is 1:1:

1. **Training cohort**: 2,688 subjects (3,263 scans) were assigned to training cohort which contained 1,230 non-pneumonia, 666 CAP, 41 influenza-A/B and 751 COVID-19 subjects. In this cohort, 198 CAP subjects (198 scans) and 468 COVID-19 subjects (725 scans) had localizer scans which were considered as paired CXR and the CXRs were used to train CXR diagnosis network.
2. **Test cohort:** 2,690 subjects (3,203 scans) were assigned to test cohort which contained 1,229 non-pneumonia subjects, 668 CAP, 42 influenza-A/B and 751 COVID-19 subjects. In this cohort, 220 CAP subjects (220 scans) and 469 COVID-19 subjects (802 scans) had localizer scans which were considered as paired CXR. This part of data was used to evaluate CT performance compared with CXR.

CC-CCII database shares only the image slices and some unknown processes had been performed to extract those slices from CT volumes. Besides, some of their slices were raw image slices and some others were cropped by lung masks. Its category definition is also a little different from ours. Due to these reasons, we trained and evaluated our system separately on CC-CCII database. We divided all its 2,539 subjects (3,784 scans) into training and test cohort with rate of 1:1, resulting in 1,269 subjects (1,841 scans) in **CC-CCII training cohort** and 1,270 subjects (1,943 scans) in **CC-CCII test cohort**.

### Development of Deep Learning Modules

The lung segmentation module is implemented based on U-Net^34^, which is a 2D semantic segmentation network. All CTs are in 3D, so we trained and tested the segmentation model slice by slice. The training slices were extracted from chest CTs in the training cohort and annotations of lung segmentation were obtained manually. Lung segmentations work as masks and region boxes in diagnosis module (detailly described in **supplementary methods**).

The slice diagnosis module is a 2D classification deep network whose backbone is ResNet152^35^. The parameters of ResNet152 are pretrained on a huge dataset ImageNet^9^ for better and faster convergence. We tested a 3D classification network but this 2D scheme showed much better performance and 3D network might not work if GPU memory is limited (even 11G memory might not be able to process one volume). In order to make sure different scanners and factors other than lung area will not influence the diagnosis, the inputs of slice diagnosis module were lung-masked slices which have been cropped out along their lung segmentations’ bounding boxes and the segmentations were also input as another channel. The outputs of module were four scores respectively representing confidence levels of being four categories (three categories for CC-CCII database). Slices for training this network were extracted from training scans, and the extraction process is detailly explained in supplementary methods.

A task-specific fusion block is used to get a volume/case-level prediction from slice-level results. Because one volume is regarded as pneumonia infected if any one of its slices is diagnosed as infected, the fused scores of three pneumonia classes (CAP, Influenza, and COVID-19) were obtained by averaging the top-*K* highest scores of each class (*K* was 3 in our experiments) in all slices and the non-pneumonia score is obtained by averaging all slices. If the task is specific to distinguish non-pneumonia and pneumonia subjects, all pneumonia scores of a slice will be summed up. If the task is specific to distinguish COVID-19 from other pneumonia, the scores of other classes will be muted in the fusion block. To measure the diagnostic performance, AUC, sensitivity and specificity (with default threshold 0.5) are computed on test cohort.

COVID-infectious slice locating module has the same structure as the slice diagnosis module but it was trained and evaluated especially on a set of COVID-19 positive subjects whose slices with lesions have been marked manually. All training samples of this block came from training cohort, and evaluation samples came from test cohort.

We used Guided Grad-CAM to obtain attentional regions as our system interpreting block. Guided Grad-CAM has the advantage that it not only generates a heat map to locate the relevant area, but also produces a coarse localization map highlighting the important regions in the image for predicting the result. Guided Grad-CAM is important because the areas it focused on are secondary outputs of our system together with diagnosis result, giving more detailed diagnosis suggestions. Also, the attentional regions were used in latter feature extraction and analysis to get more detailed information about lesion areas. We extracted region of attention by binarizing the output of Guided Grad-CAM and then some morphological operations were done on the binarization map.

T-SNE was done based on features from slice diagnosis module, which were features before the last fully-connected layer of every slices of a volume, and max-pooling them on dimension of slices. Therefore, every CT volume was mapped to a 2048-d latent which was used to perform t-SNE.

All the deep learning blocks were implemented using *PyTorch*^36^. T-SNE was implemented using *“scikit-learn”* package.

### Reader Study

In order to detailly analyze the performances, we set up three different reader study cohorts for different diagnosis tasks, which were acquired by randomly selecting subjects from test cohort:

1. **Pneumonia-or-non-pneumonia cohort**: 100 subjects (50 non-pneumonia subjects, 25 CAP subjects and 25 COVID-19 subjects from three centers) were assigned. This cohort was used to compare results of the AI system with radiologists in diagnosis of pneumonia.
2. **CAP-or-COVID-19 cohort**: 100 subjects (50 CAP subjects and 50 COVID-19 subjects from three centers) were assigned. This cohort was used to compare results of the AI system with radiologists in distinguishing COVID-19 from CAP.
3. **Influenza-or-COVID-19 cohort**: 50 subjects (20 influenza-A/B subjects and 30 COVID-19 subjects from three centers) were assigned. This cohort was used to compare results of the AI system with radiologists in distinguishing COVID-19 from influenza-A/B.

We invited five professional radiologists in our experiments who are all radiologists in the radiology department of Wuhan Union Hospital and have rich clinical diagnosis experience and is in the center of the epidemic area with the most patients in this outbreak in China. They have all read over four-hundred CTs of COVID-19 in the past three months. Five radiologists had an average of 8 years of clinical experience in the imaging diagnosis of pulmonary diseases, as detailed in **Extended Data Table 3 A**.

The human radiologists were aware of the tasks and the possible classes in reader cohort when reading. For example, they were informed that only CAP and COVID-19 subjects were collected in CAP-or-COVID cohort. Besides, readers can choose any window of gray value and zoom in or out when reading CT volumes using *Slicer 4.10.2* software while our system used fixed size resample images (224 × 224 × 35) with fixed gray value window (−1200, 700) for all volumes.

### Attentional Region Extraction

To analyze the differences of imaging phenotype between different pneumonia, features were extracted in the attentional region determined by Guided Grad-CAM. The attentional regions were determined by binarizing Grad-CAM maps and operating morphological transforms. We only kept the regions with valid size (larger than 200 pixels within margin of lung mask). We extracted attentional regions only in pneumonia subjects in test cohort.

### Phenotype Feature Extraction

Radiomics features widely used in tumor diagnosis were extracted. These features were composed of different image transforms and feature matrix calculations. We adopted three image transforms: original image, transformed image by Laplacian of Gaussian (LoG) operator, and transformed image by wavelet. For each image after the operation of a transform, six series of features are extracted, including first order features, Gray Level Co-occurrence Matrix (GLCM), Gray Level Size Zone Matrix (GLSZM), Gray Level Run Length Matrix (GLRLM), Neighboring Gray Tone Difference Matrix (NGTDM), Gray Level Dependence Matrix (GLDM). Radiomics analysis was performed using *python version 3.6* and the “*pyradiomics*” package^30^.

The distance feature was defined as the distance between the center of gravity of the region of interest (obtained by Grad-CAM) and the edge of the lung (obtained by lung segmentation results). Besides, 2D contour fractal dimension and 3D grayscale mesh fractal dimension of the attentional region were extracted. The fractal dimension describes the degree of curvature of a curve or surface.

LASSO logistic regression model was used to choose most discriminative features in all extracted ones. LASSO analysis was performed using *python version 3.6* and the “ *scikit-learn”* package.

### Phenotype Feature Analysis

P-value of selected radiomics features and the sum of them weighted by their coefficients were calculated (**Extended Data Figure 4, 5**). We found that all features selected to distinguish CAP and COVID-19 was significant in t-test. KS test is used to test whether two groups of value come from two distributions, according to which *wavelet-LL_firstorder_10Percentile* and *diagnostics_Image-original_Mean* can help divide CAP and COVID-19 into two distributions. However, results for distinguishing influenza-A/B and COVID-19 were all not significant. We believed that radiomics features and LASSO model might have some limitation in such unbalanced dataset (only 73 influenza-A/Bs but 1,204 scans for COVID-19). Our deep learning-based diagnosis model can however overcome this limitation.

Distance feature was defined as the distances of attentional regions of all three types of pneumonia to lung boundaries (**Extended Data Figure 6 A**). There were two peaks of distances of COVID-19 which were generally 0-30 pixels (2.5 mm/pixel) from the pleura and a little amount of which were larger than 100 pixels. That is different with other distributions that most of CAPs were not larger than 40 pixels to pleura. Distribution of influenza-A/B was flatter, which is consistent with anatomical findings on COVID-19. We found that the distribution was consistent with pathological study. When the SARS-Cov2 virus is inhaled through the airways, it mainly invades the deep bronchioles, causes inflammation of the bronchioles and their surroundings to damage alveolar^37,38^. These areas have well-established immune system and well-developed pulmonary lobules, leading to a strong inflammatory response^39,40^. Secondly, due to region of attention obtained by Grad-CAM does not delineate the lesion accurately, there were little differences among three types of pneumonia in fractal dimension (**Extended Data Figure 6 B**). Thirdly, COVID-19 was a little fickler in gray-value compared to others with higher 3D fractal dimension, while CAP had two peaks in this feature. According to p-value, distance feature and 3D fractal dimension can help distinguish CAP and COVID-19, the 2D fractal dimension showed little information (**Extended Data Figure 6 C**). No significant difference could be found between influenza-A/B and COVID-19 on those features and according to KS test only distance feature is significative to distinguish CAP and COVID-19.

According to all selected features, we can describe in depth the relationship between the medical findings and typical patterns of COVID-19 (some examples are shown in **supplementary Figure 5**). I) Halo and anti-halo pattern. The halo pattern was speculated to be that the lesions (mainly the central node of the lobular) infiltrates into the surrounding interstitium and developed the aggregation of inflammatory cells in the interstitium. Anti-halo pattern was ground glass shadow surrounded by the high-density consolidation. The appearance of this sign may be that the inflammatory repair was dominated by the edge, leading to the formation of a band shadow tending to consolidation at the edge, while the central repair was relatively slow. II) Pleural parallel signs. The formation mechanism was speculated as follows: when the COVID-19 invaded the interstitium around the alveoli, the lymphatic return direction was subpleural and interlobular septa, and diffused into pleural side and bilateral interlobular septum^41^. Because of the limitation of the pleura at the distal end, the lymph can only cling to the pleura and spread along the reticular structure of the interlobular septal margin on both sides. In addition, the fusion of the subpleural lesions resulted in the long axis of the lesions parallel to the pleura. III) Vascular thickening. It was consistent with the rules of inflammation production. The inflammatory increased vascular permeability, caused telangiectasia, further caused pulmonary artery thickening^40,42^. IV) The fine mesh feature of large area. The COVID-19 mainly invaded the interstitium in the lobules, so it appeared as confluent fine mesh (crazy paving). V) The density-increased GGO. This kind of GGO was transforming to consolidation. The consolidation edges were flat or contracted, and fiber strands appeared. Compared with COVID-19, influenza showed higher density on the lesion area, which can be inferred that they caused slightly less alveolitis and GGO patterns. As a result of bronchiolitis, influenza virus pneumonia was more likely to form tree-bud signs, and occasionally hyperlucent lung.

---

**Extended Data Table 1.** Characteristics of patients from Wuhan Union Hospital, Western Campus of Wuhan Union Hospital, Jianghan Mobile Cabin Hospital.

**A**
| # Subject ( # Scan) | Non-pneumonia | CAP | Influenza-A/B | COVID-19 | In total |
| --- | --- | --- | --- | --- | --- |
| Training cohort | 1230 (1233) | 666(666) | 41 (70) | 751 (1294) | 2688 (3263) |
| Test cohort | 1229 (1234) | 666 (668) | 42 (62) | 751 (1235) | 2690 (3203) |
| CC-CCII training cohort | 424 (539) | 482 (726) | 0 (0) | 363 (576) | 1269 (1841) |
| CC-CCII test cohort | 425 (540) | 482 (822) | 0 (0) | 363 (681) | 1270 (1943) |
| In total | 3308 (3546) | 2298 (2886) | 83 (132) | 2228 (3686) | 7917 (10250) |

**B**
| # Subject |  | Non-pneumonia |  | CAP |  | Influenza-A/B |  | COVID-19 |  |
| --- | --- | --- | --- | --- | --- | --- | --- | --- | --- |
|  |  | Training Cohort | Test Cohort | Training Cohort | Test Cohort | Training Cohort | Test Cohort | Training Cohort | Test Cohort |
| Gender | male | 25 | 34 | 404 | 424 | 27 | 27 | 348 | 369 |
|  | female | 104 | 95 | 262 | 244 | 14 | 15 | 403 | 382 |
| Age | <=20 | 0 | 1 | 62 | 69 | 2 | 0 | 1 | 3 |
|  | 21-40 | 122 | 118 | 116 | 98 | 5 | 7 | 139 | 116 |
|  | 41-60 | 6 | 10 | 193 | 198 | 7 | 8 | 268 | 261 |
|  | 61-80 | 1 | 0 | 241 | 243 | 19 | 20 | 314 | 335 |
|  | >80 | 0 | 0 | 45 | 50 | 8 | 7 | 29 | 36 |
|  | no-record | 0 | 0 | 9 | 10 | 0 | 0 | 0 | 0 |
| # Stage | 1 | 129 | 129 | 666 | 664 | 28 | 27 | 323 | 341 |
|  | 2 | 0 | 0 | 0 | 4 | 7 | 11 | 300 | 301 |
|  | 3 | 0 | 0 | 0 | 0 | 1 | 1 | 103 | 87 |
|  | >3 | 0 | 0 | 0 | 0 | 5 | 3 | 25 | 22 |

**A.** Number of each classes computed by subjects (persons) and scans. **B.** Detailed characteristics of subjects of training and test cohort from Wuhan. **Note:** The characteristics of publicly available databases were cleared after anonymization, so Table B only lists the data from Wuhan.

**Extended Data Table 2.** Metrics of the proposed AI system.

| Task | AUC (95% CI) | Sensitivity (95% CI) | Specificity (95% CI) |
| --- | --- | --- | --- |
| Non-pneumonia diagnosis | 0.9752<br>(0.9726-0.9783) | 0.9343<br>(0.9290-0.9429) | 0.9801<br>(0.9778-0.9827) |
| CAP diagnosis | 0.9804<br>(0.9776-0.9837) | 0.9687<br>(0.9634-0.9741) | 0.9407<br>(0.9366-0.9448) |
| Influenza-A/B diagnosis | 0.9885<br>(0.9861-0.9928) | 0.8307<br>(0.7962-0.8696) | 0.9945<br>(0.9936-0.9960) |
| COVID-19 diagnosis | 0.9745<br>(0.9722-0.9771) | 0.8703<br>(0.8620-0.8784) | 0.9660<br>(0.9629-0.9693) |
| COVID-infectious slices locating | 0.9559<br>(0.9532-0.9586) | 0.8009<br>(0.79323-0.8094) | 0.9636<br>(0.9607-0.9666) |

B
| Task | AUC (95% CI) | Sensitivity (95% CI) | Specificity (95% CI) |
| --- | --- | --- | --- |
| Normal diagnosis | 0.9881<br>(0.98634-0.9901) | 0.9814<br>(0.9771-0.9862) | 0.95381<br>(0.9487-0.9590) |
| Common pneumonia diagnosis | 0.9907<br>(0.9895-0.9922) | 0.9282<br>(0.9210-0.9371) | 0.9706<br>(0.9664-0.9753) |
| COVID-19 diagnosis | 0.9778<br>(0.9750-0.9808) | 0.9277<br>(0.9197-0.9376) | 0.9903<br>(0.9881-0.9926) |

C
| Task | AUC (95% CI) | Sensitivity (95% CI) | Specificity (95% CI) |
| --- | --- | --- | --- |
| CT COVID-19 diagnosis | 0.9847<br>(0.9822-0.9877) | 0.9762<br>(0.9718-0.98110) | 0.91250<br>(0.8975-0.9301) |
| CXR COVID-19 diagnosis | 0.9527<br>(0.9474-0.9583) | 0.9623<br>(0.9570-0.9673) | 0.7155<br>(0.6918-0.7436) |
| Result-level fusion | 0.9894<br>(0.9873-0.9917) | 0.9469<br>(0.9399-0.9543) | 0.9503<br>(0.9371-0.9627) |

D
| Task | AUC (95% CI) | Sensitivity (95% CI) | Specificity (95% CI) |
| --- | --- | --- | --- |
| Pneumonia-or-non-pneumonia cohort | 0.9869<br>(0.9818-0.9993) | 0.9416<br>(0.9210-0.9750) | 0.9203<br>(0.8974-0.9524) |
| CAP-or-COVID-19 cohort | 0.9727<br>(0.9637-0.9825) | 0.9591<br>(0.9459-0.9767) | 0.9199<br>(0.8947-0.9512) |
| Influenza-or-COVID-19 cohort | 0.9585<br>(0.9413-0.9813) | 0.94961<br>(0.93333-1.0) | 0.8331<br>(0.7826-0.8846) |

**A.** Metrics on test cohort and COVID-infectious slices locating. **B.** Metrics on CC-CCII test cohort. **C.** Metrics on comparison between CT and XCT on paired data of test cohort. **D.** Metrics on three reader study tasks.

**Extended Data Figure 1.**
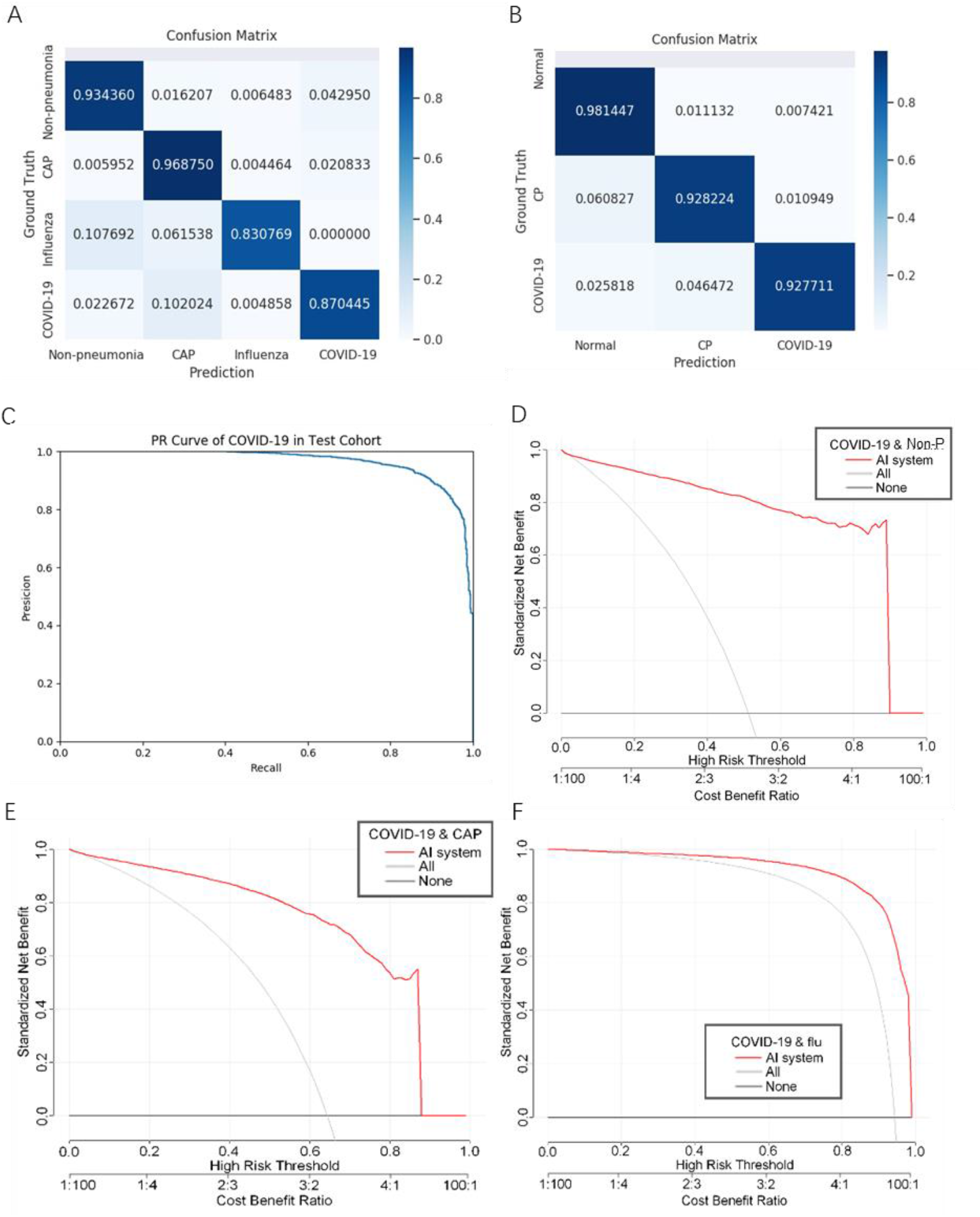
Performances of the proposed AI system. **A**. Confusion matrix of diagnosis model on test cohort. **B**. Confusion matrix of diagnosis model on CC-CCII test cohort **C**. PR curves was employed to assess the AI system performance of COVID-19 diagnosis. **D-F**. Decision curve analyses (DCA) of the AI system on test set for diagnosis respectively between COVID-19 and non-pneumonia, CAP and influenza-A/B.

**Extended Data Table 3.** Reader study statistics and results.

| Reader information |  |  |  |
| --- | --- | --- | --- |
|  | Reads per year | Years of experience | Reads on COVID-19 |
| Reader 1 | 3000-4000 | 5 | 500-600 |
| Reader 2 | 4000-5000 | 10 | 500-600 |
| Reader 3 | 4000-5000 | 11 | 600-700 |
| Reader 4 | 3000-4000 | 8 | 600-700 |
| Reader 5 | 3000-4000 | 7 | 500-600 |

B
|  |  |  | All Reader |  | Any Reader |  |
| --- | --- | --- | --- | --- | --- | --- |
|  |  |  | Correct | Wrong | Correct | Wrong |
| AI | Pneumonia-or-non-pneumonia | Correct | 97 | 0 | 95 | 2 |
|  |  | Wrong | 2 | 1 | 2 | 1 |
|  | CAP-or-COVID | Correct | 92 | 2 | 69 | 23 |
|  |  | Wrong | 5 | 1 | 5 | 3 |
|  | Influenza -or-COVID | Correct | 44 | 3 | 24 | 20 |
|  |  | Wrong | 3 | 0 | 3 | 3 |

C
|  |  |  | Reader 1 |  | Reader 2 |  | Reader 3 |  | Reader 4 |  | Reader 5 |  |
| --- | --- | --- | --- | --- | --- | --- | --- | --- | --- | --- | --- | --- |
|  |  |  | C | W | C | W | C | W | C | W | C | W |
| AI | Pneumonia-or-non-pneumonia | C | 96 | 1 | 96 | 1 | 96 | 1 | 96 | 1 | 96 | 1 |
|  |  | W | 2 | 1 | 2 | 1 | 2 | 1 | 2 | 1 | 2 | 1 |
|  | CAP-or-COVID | C | 86 | 7 | 81 | 12 | 85 | 8 | 82 | 11 | 80 | 13 |
|  |  | W | 6 | 1 | 4 | 3 | 6 | 1 | 5 | 2 | 5 | 2 |
|  | Influenza-or-COVID | C | 33 | 11 | 31 | 13 | 34 | 10 | 35 | 9 | 35 | 9 |
|  |  | W | 5 | 1 | 3 | 3 | 5 | 1 | 5 | 1 | 4 | 2 |

**A.** Experience levels of the five radiologists involved in the reader study. **B.** Comparison of diagnostic error between the AI system and human readers. **Notes:** In **Any Reader** column, readers are deemed as wrong if anyone is wrong, otherwise deemed as correct. In **All Reader** column, readers are deemed as wrong if all were wrong, otherwise deemed as correct. All AI diagnosis was working with 0.5 threshold. **C.** Comparison of diagnostic performances between the AI system and every human reader. **Notes:** **C** means correct and **W** means wrong.

**Extended Data Figure 2.**
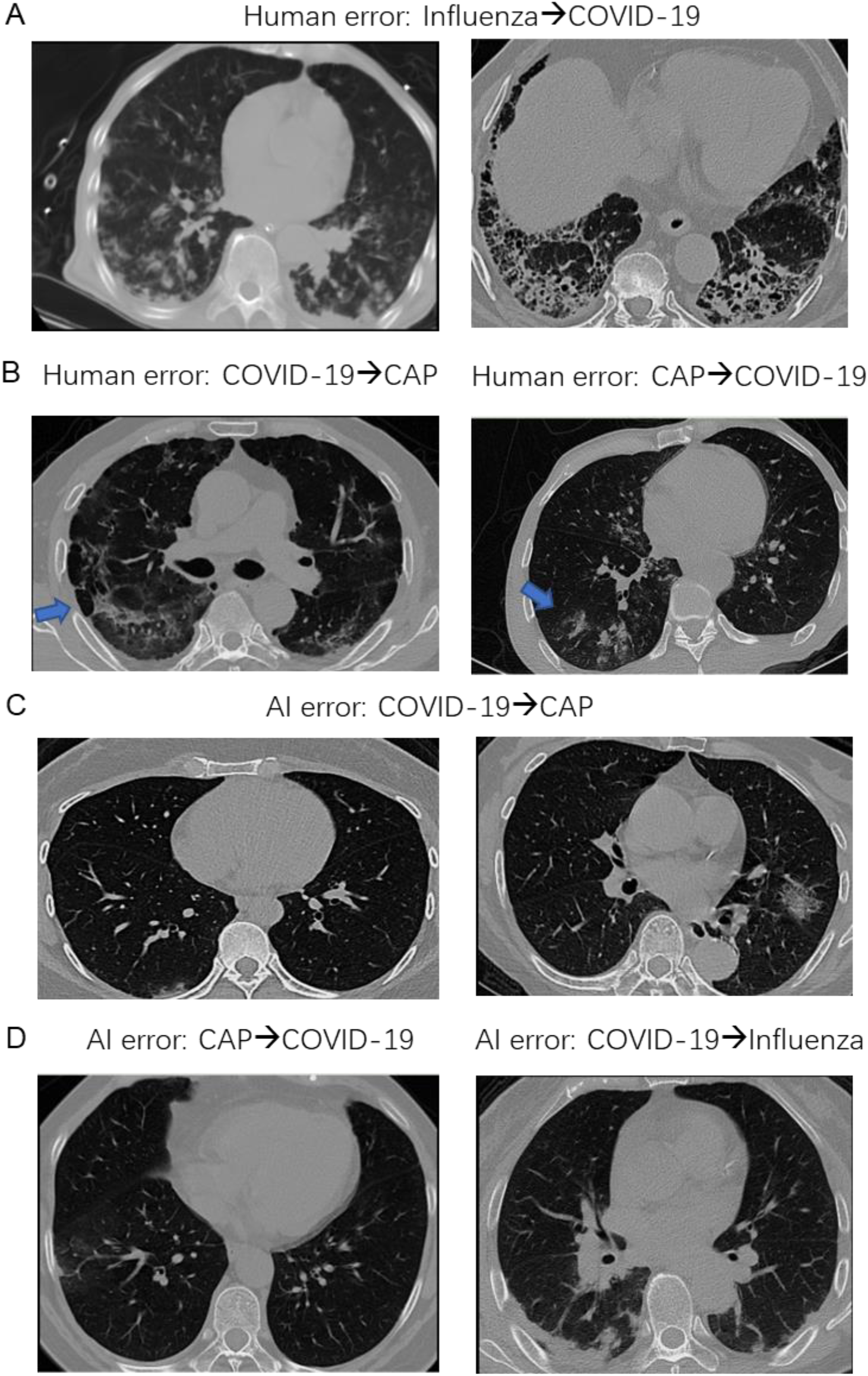
Examples for wrong classification by human readers and the AI system. **A**. Two influenza samples that all five readers misclassified. **B**. Two samples that all five readers misclassified. The left one is CAP cases and the right one is COVID-19 cases. **C**. Two COVID-19 samples that were diagnosed as CAP by AI. **D**. Two samples AI diagnosed wrongly. The left one is CAP and the right one is COVID-19.

**Extended Data Figure 3.**
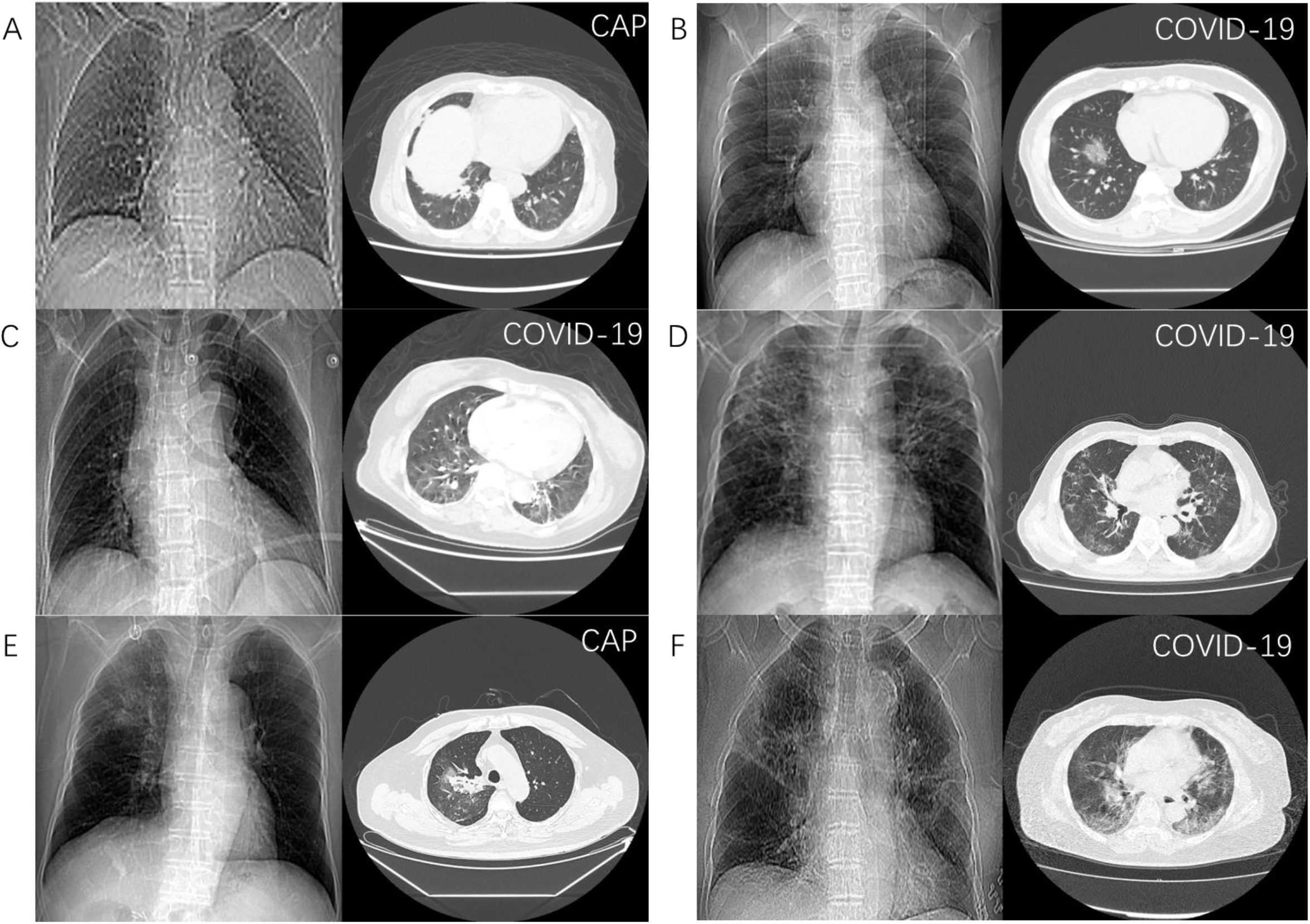
Representative diagnosis results of CT-based AI system and CXR-based AI system. **A**. An example that both CXR and CT were wrong. **B**. An example that both CXR and CT were correct. **C**. An example that CXR was correct, CT was wrong, and fusion result was wrong. **D**. An example that CT was correct, CXR was wrong, and fusion result was correct. E. An example that CXR was correct, CT was wrong, and fusion was correct. F. A case that CXR was wrong, CT was correct, and fusion was wrong. **Note:** the true classes were annotated at the upper right of every images.

**Extended Data Figure 4.**
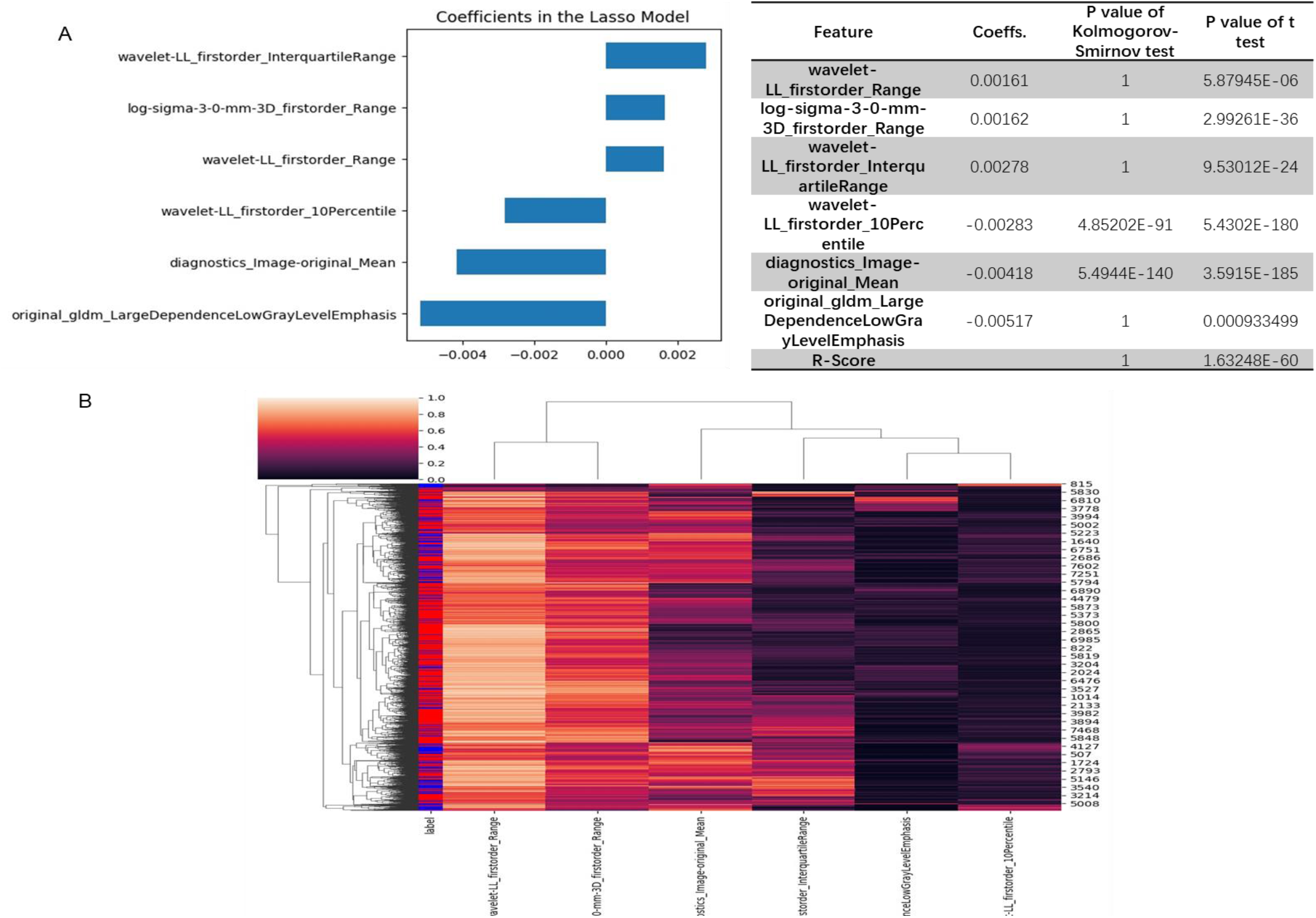
Selected radiomics features to identify COVID-19 from CAP. **A**. Coefficients of selected features and the R-score provided by LASSO model based on those features. **B**. Cluster heatmap of the selected features.

**Extended Data Figure 5.**
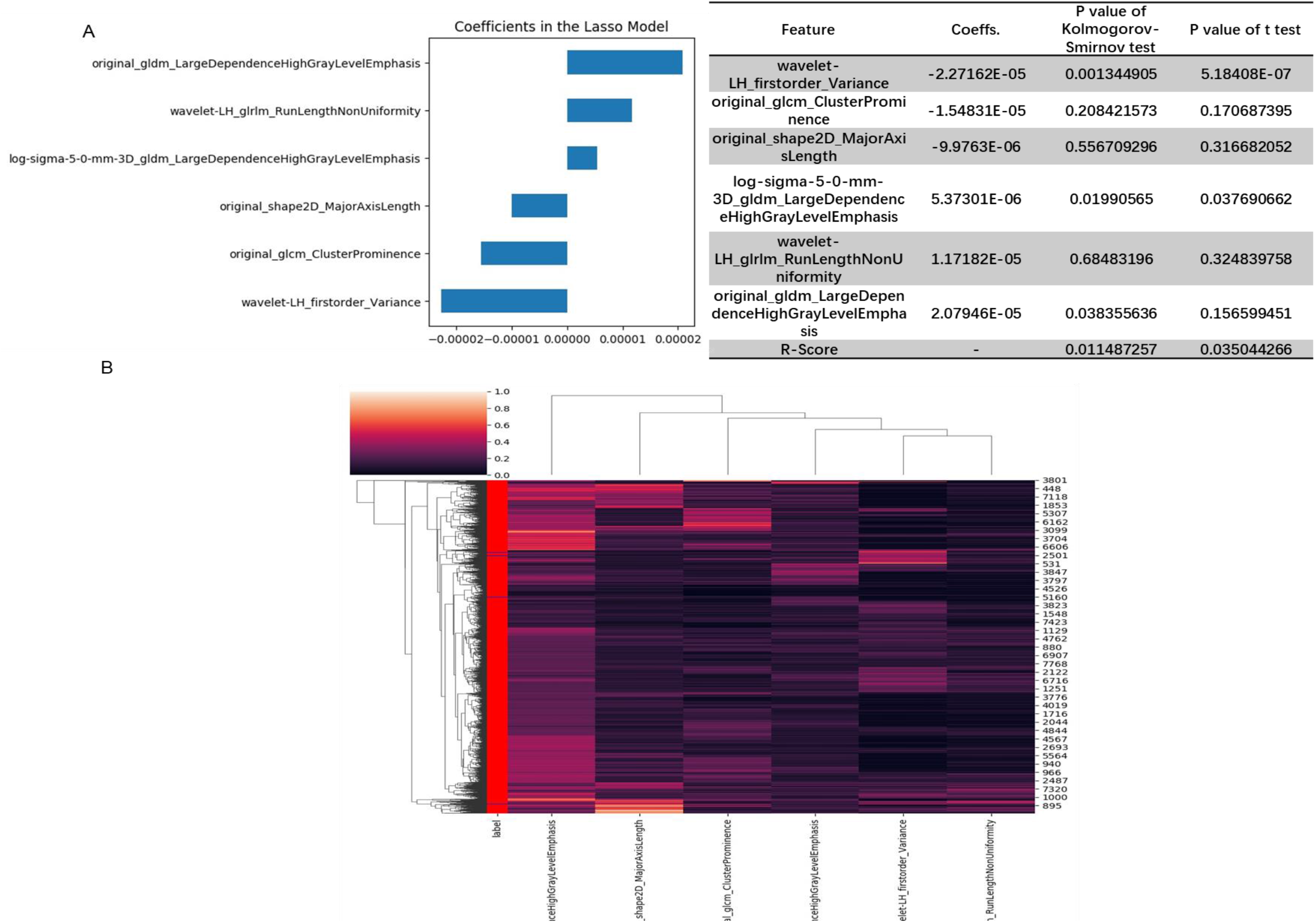
Selected radiomics features to identify COVID-19 from influenza-A/B. **A**. Coefficients of selected features and the R-score provided by LASSO model based on those features. **B**. Cluster heatmap of the selected features.

**Extended Data Figure 6.**
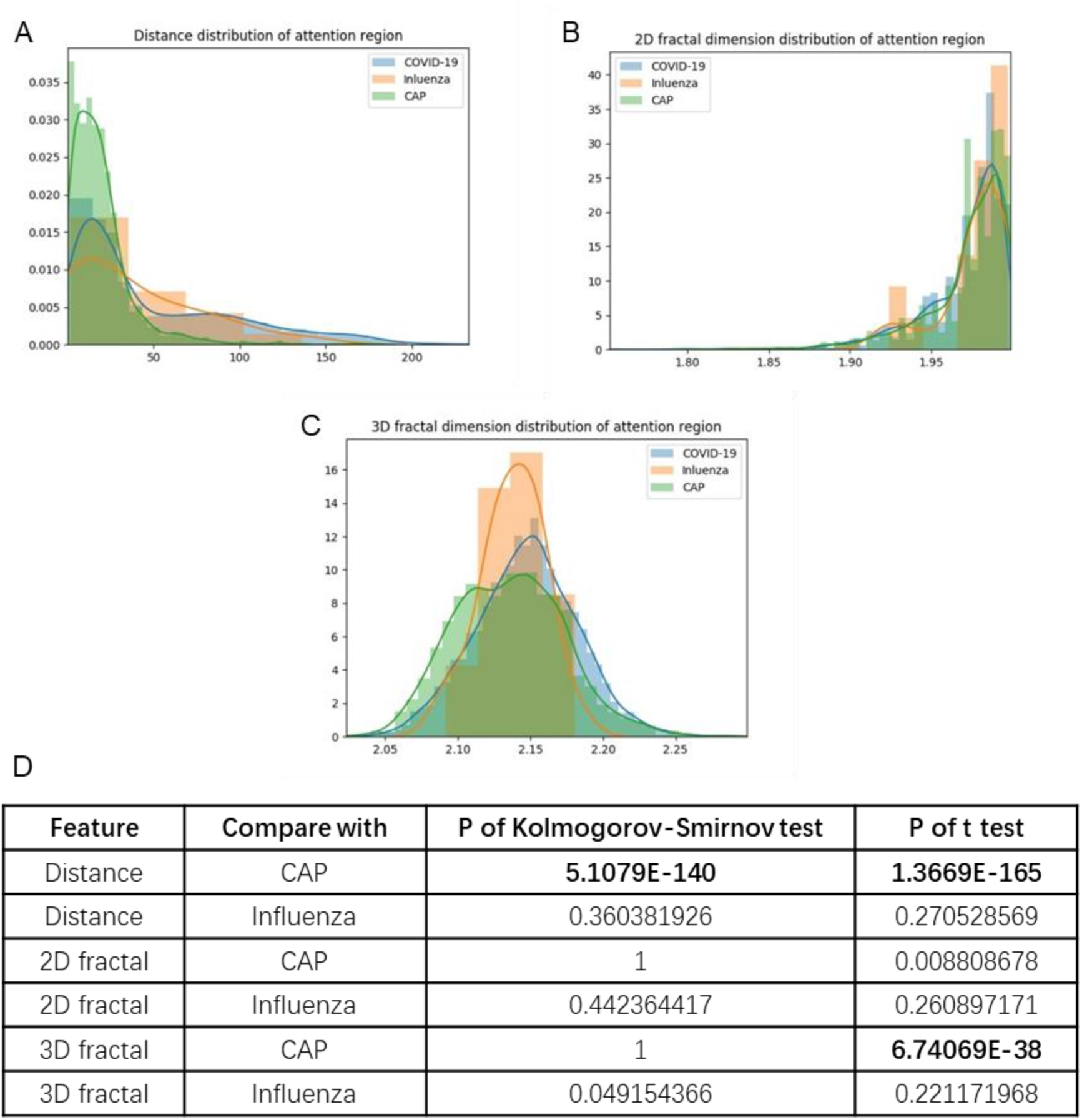
Distribution of three attentional region related features. **A**. Distances from the center of attentional region to lung’s margin. **B**. Margin fractal dimension of attentional region. **C**. Gray level mesh fractal dimension off attentional region. **D**. p value of Kolmogorov-Smirnov test and t test of three features between COVID-19 and CAP or influenza.

## Reference

1 Laboratory testing for coronavirus disease 2019 (COVID-19) in suspected human cases: interim guidance, 2 March 2020. (World Health Organization, 2020).

2 Bai, H. X. et al. Performance of radiologists in differentiating COVID-19 from viral pneumonia on chest CT. Radiology, 200823 (2020).

3 Ai, T. et al. Correlation of chest CT and RT-PCR testing in coronavirus disease 2019 (COVID-19) in China: a report of 1014 cases. Radiology, 200642 (2020).

4 Rubin, G. D. et al. The role of chest imaging in patient management during the COVID-19 pandemic: a multinational consensus statement from the Fleischner Society. Chest (2020).

5 Wong, H. Y. F. et al. Frequency and distribution of chest radiographic findings in COVID-19 positive patients. Radiology, 201160 (2020).

6 Shi, H. et al. Radiological findings from 81 patients with COVID-19 pneumonia in Wuhan, China: a descriptive study. The Lancet Infectious Diseases (2020).

7 Deng, J. et al. Imagenet: A large-scale hierarchical image database. in 2009 IEEE conference on computer vision and pattern recognition. 248–255 (IEEE, 2009 Published).

8 LeCun, Y., Bengio, Y. & Hinton, G. Deep learning. Nature 521, 436–444 (2015).

9 Krizhevsky, A., Sutskever, I. & Hinton, G. E. Imagenet classification with deep convolutional neural networks. in Advances in neural information processing systems. 1097-1105 (2012 Published).

10 Ren, S., He, K., Girshick, R. & Sun, J. Faster r-cnn: Towards real-time object detection with region proposal networks. in Advances in neural information processing systems. 91–99 (2015 Published).

11 Esteva, A. et al. Dermatologist-level classification of skin cancer with deep neural networks. Nature 542, 115–118 (2017).

12 Litjens, G. et al. A survey on deep learning in medical image analysis. Medical image analysis 42, 60–88 (2017).

13 Esteva, A. et al. A guide to deep learning in healthcare. Nature medicine 25, 24–29 (2019).

14 Topol, E. J. High-performance medicine: the convergence of human and artificial intelligence. Nature medicine 25, 44–56 (2019).

15 Ardila, D. et al. End-to-end lung cancer screening with three-dimensional deep learning on low-dose chest computed tomography. Nature medicine 25, 954–961 (2019).

16 Shi, F. et al. Review of artificial intelligence techniques in imaging data acquisition, segmentation and diagnosis for covid-19. IEEE Reviews in Biomedical Engineering (2020).

17 Dong, D. et al. The role of imaging in the detection and management of COVID-19: a review. IEEE Reviews in Biomedical Engineering (2020).

18 Zhang, K. et al. Clinically Applicable AI System for Accurate Diagnosis, Quantitative Measurements and Prognosis of COVID-19 Pneumonia Using Computed Tomography. Cell (2020).

19 Li, L. et al. Artificial Intelligence Distinguishes COVID-19 from Community Acquired Pneumonia on Chest CT. Radiology, 200905, doi:10.1148/radiol.2020200905 (2020).

20 Mei, X. et al. Artificial intelligence–enabled rapid diagnosis of patients with COVID-19. Nature Medicine, 1-5 (2020).

21 Bai, H. X. et al. AI augmentation of radiologist performance in distinguishing COVID-19 from pneumonia of other etiology on chest CT. Radiology, 201491 (2020).

22 Wang, X. et al. A Weakly-supervised Framework for COVID-19 Classification and Lesion Localization from Chest CT. IEEE Transactions on Medical Imaging, doi:10.1109/TMI.2020.2995965 (2020).

23 Han, Z. et al. Accurate Screening of COVID-19 using Attention Based Deep 3D Multiple Instance Learning. IEEE Transactions on Medical Imaging, doi:10.1109/TMI.2020.2996256 (2020).

24 Ouyang, X. et al. Dual-Sampling Attention Network for Diagnosis of COVID-19 from Community Acquired Pneumonia. IEEE Transactions on Medical Imaging, doi:10.1109/TMI.2020.2995508 (2020).

25 Murphy, K. et al. COVID-19 on the Chest Radiograph: A Multi-Reader Evaluation of an AI System. Radiology, 201874 (2020).

26 Armato, S. G., 3rd et al. The Lung Image Database Consortium (LIDC) and Image Database Resource Initiative (IDRI): a completed reference database of lung nodules on CT scans. Medical physics 38, 915–931, doi:10.1118/1.3528204 (2011).

27 Tianchi Competition, <https://tianchi.aliyun.com/competition/entrance/231601/information> (2017).

28 Selvaraju, R. R. et al. Grad-cam: Visual explanations from deep networks via gradient-based localization. in Proceedings of the IEEE International conference on computer vision. 618–626 (IEEE, 2017 Published).

29 Maaten, L. v. d. & Hinton, G. Visualizing data using t-SNE. Journal of machine learning research 9, 2579-2605 (2008).

30 Van Griethuysen, J. J. et al. Computational radiomics system to decode the radiographic phenotype. Cancer research 77, e104-e107 (2017).

31 Bernheim, A. et al. Chest CT findings in coronavirus disease-19 (COVID-19): relationship to duration of infection. Radiology, 200463 (2020).

32 Xiong, Q. et al. Women may play a more important role in the transmission of the corona virus disease (COVID-19) than men. Lancet (2020).

33 Kanne, J. P. Chest CT findings in 2019 novel coronavirus (2019-nCoV) infections from Wuhan, China: key points for the radiologist. Report No. 0033–8419, 16-17 (Radiological Society of North America, 2020).

## References

34 Ronneberger, O., Fischer, P. & Brox, T. U-net: Convolutional networks for biomedical image segmentation. in International Conference on Medical image computing and computer-assisted intervention. 234–241 (Springer, 2015 Published).

35 He, K., Zhang, X., Ren, S. & Sun, J. Deep residual learning for image recognition. in Proceedings of the IEEE conference on computer vision and pattern recognition. 770–778 (IEEE, 2016 Published).

36 Paszke, A. et al. PyTorch: An imperative style, high-performance deep learning library. in Advances in Neural Information Processing Systems. 8024-8035 (2019 Published).

37 Cascella, M., Rajnik, M., Cuomo, A., Dulebohn, S. C. & Di Napoli, R. Features, evaluation and treatment coronavlrus (COVID-19). in StatPearls [Internet], (StatPearls Publishing, 2020).

38 Dail, D. H., & Hammar, S. P. Dall and Hammar’s pulmonary pathology. (Springer Science & Business Media, 2013).

39 Cotes, J. E., Chinn, D. J. & Miller, M. R. Lung function: physiology, measurement and application in medicine. (John Wiley & Sons, 2009).

40 Panagiotou, M., Church, A. C., Johnson, M. K. & Peacock, A. J. Pulmonary vascular and cardiac impairment in interstitial lung disease. Eur Respir Rev 26, doi:10.1183/16000617.0053-2016 (2017).

41 Breslin, J. W. et al. Lymphatic vessel network structure and physiology. Comprehensive Physiology 9, 207–299 (2011).

42 Moldoveanu, B. et al. Inflammatory mechanisms in the lung. Journal of inflammation research 2, 1 (2009).

